# Evaluating ChatGPT as an Adjunct for Radiologic Decision-Making

**DOI:** 10.1101/2023.02.02.23285399

**Authors:** Arya Rao, John Kim, Meghana Kamineni, Michael Pang, Winston Lie, Marc D. Succi

**Author notes:** **Corresponding Author:** Marc D. Succi, MD, Massachusetts General Hospital, Department of Radiology, 55 Fruit Street, Boston, MA, 02114, @MarcSucciMD.

## Abstract

**BACKGROUND:** ChatGPT, a popular new large language model (LLM) built by OpenAI, has shown impressive performance in a number of specialized applications. Despite the rising popularity and performance of AI, studies evaluating the use of LLMs for clinical decision support are lacking.

**PURPOSE:** To evaluate ChatGPT’s capacity for clinical decision support in radiology via the identification of appropriate imaging services for two important clinical presentations: breast cancer screening and breast pain.

**MATERIALS AND METHODS:** We compared ChatGPT’s responses to the American College of Radiology (ACR) Appropriateness Criteria for breast pain and breast cancer screening. Our prompt formats included an open-ended (OE) format, where ChatGPT was asked to provide the single most appropriate imaging procedure, and a select all that apply (SATA) format, where ChatGPT was given a list of imaging modalities to assess. Scoring criteria evaluated whether proposed imaging modalities were in accordance with ACR guidelines.

**RESULTS:** ChatGPT achieved an average OE score of 1.83 (out of 2) and a SATA average percentage correct of 88.9% for breast cancer screening prompts, and an average OE score of 1.125 (out of 2) and a SATA average percentage correct of 58.3% for breast pain prompts.

**CONCLUSION:** Our results demonstrate the feasibility of using ChatGPT for radiologic decision making, with the potential to improve clinical workflow and responsible use of radiology services.

## INTRODUCTION

Many artificial intelligence models have been utilized in healthcare to aid in clinical decision support (CDS), including for applications in radiology. Yet, use cases are mostly limited to imaging interpretation-- in radiology, this includes identifying relevant image features and summarizing non-image patient data ^1^. Other applications of AI in radiology focus on medical education, including providing real-time feedback to radiology trainees when providing diagnoses with radiology images ^2^. In this study, we propose and evaluate a novel use of artificial intelligence in radiologic CDS: identifying appropriate imaging services based on an initial clinical presentation.

ChatGPT is a large language model (LLM) built on Generative Pre-trained Transformer 3.5 (GPT-3.5), fine-tuned with human supervision and reinforcement ^3^. Since its public release in November 2022, the chatbot from OpenAI has allowed over a million users to access and dialogue with its deep-learning, autoregressive LLM whose precursor (GPT-3) had at least 175 billion parameters under its training ^4^. Despite several constraints, recent reports have shown promising ChatGPT performance in the MBA degree exam ^5^, the United States Certified Public Accountant exam ^6^, the Uniform Bar exam ^7^, and the United States Medical Licensing Exam ^8^ demonstrating its ability to generate accurate textual responses to complex input criteria. ChatGPT’s ability to simulate highly technical language has prompted researchers to credit the chatbot authorship in biomedical literature ^9,10^ with scientific journals scrambling to clarify publishing ethics in response ^11–13^.

Given ChatGPT’s performance in these settings, we hypothesized that ChatGPT is capable of providing CDS in triaging patients for imaging services. To evaluate the feasibility of this application, we tested ChatGPT’s accuracy in determining appropriate imaging modalities for various clinical presentations of breast cancer screening and breast pain. Breast malignancies are a leading cause of cancer mortality in women and their screening and diagnosis constitute significant imaging volume. However, current practices of screening fall short of maximizing benefits for patients and reducing waste ^14–16^. Similarly, mastalgia is a common complaint experienced by up to 70% of women in their lifetime ^17^. Although only a significant minority presenting only with breast pain are associated with cancer, many will undergo diagnostic imaging, incurring ineffective healthcare resources ^18^. To encourage responsible use of radiology services, the American College of Radiology® (ACR) has published various appropriateness criteria (AC) since 1993. Radiologists are often tasked with interpreting these guidelines, which classify patients into discrete demographic and risk groups. Integration of an AI-based tool into existing clinical workflows and systems could drastically improve efficiency, since such tools could take advantage of the wealth of information available from patient pre-test odds, diagnostic likelihood ratios, and the medical records themselves.

Here, we explore a pilot for possible utility of ChatGPT in radiologic decision making, evaluating a variety of prompting mechanisms and clinical presentations to provide substantive performance metrics on possible use cases in clinical settings.

## METHODS

### Artificial Intelligence

ChatGPT (San Francisco, OpenAI) is a transformer-based language model that can generate human-like text. ChatGPT features multiple layers of self-attention and feed-forward neural networks allowing it to capture the context and relationships between words in the input sequence. The language model is trained on diverse sources of text including websites, articles, and books but limited to knowledge from current events until 2021 ^19^. The ChatGPT model is self-contained in that it does not have the ability to search the internet when generating responses. Instead, it predicts the most likely “token” to succeed the previous one based on patterns in its training data. Therefore, it does not explicitly search through existing information or copy existing information. All ChatGPT model output was collected from the January 9, 2023 version of ChatGPT.

### ACR Criteria

The American College of Radiology (ACR) Appropriateness Criteria for (1) breast pain and (2) breast cancer screening were selected as a ground truth comparison to ChatGPT. In brief, these criteria rate the clinical utility of diagnostic modalities given representative patient presentations in the context of breast pain and cancer. The appropriateness categories are “Usually Not Appropriate,” “May be Appropriate,” and “Usually Appropriate.” Relative radiation level is also indicated for each modality on an ordinal scale; this information was not used in our study.

### Model Input

We used two prompt formats for each ACR variant input into ChatGPT:

1. Open-ended (OE) format: Without providing the ACR list of imaging modalities, we asked ChatGPT to provide the “single most appropriate imaging procedure.” This simulates how a user might actually interact with ChatGPT. Example: “For variant ‘Breast cancer screening. Average-risk women: women with <15% lifetime risk of breast cancer.’, determine the single most appropriate imaging procedure.”
2. Select All That Apply (SATA) format: We provided ChatGPT with the ACR list of imaging modalities for each variant and asked it to assess the appropriateness of each. This is in line with the actual usage of the ACR guidelines. Example: “For variant ‘Breast cancer screening. Average-risk women: women with <15% lifetime risk of breast cancer.’, assess appropriateness of the following procedures in a concise manner: Mammography screening, Digital breast tomosynthesis screening, US breast, MRI breast without and with IV contrast, MRI breast without IV contrast, FDG-PET breast dedicated, Sestamibi MBI”

All prompts are available in the Supplementary Data.

ChatGPT’s answers are informed by the context of the ongoing conversation. To avoid the influence of prior answers on model output, a new ChatGPT session was started for each prompt. To account for response-by-response variation, each prompt was tested three times, each time by a different user.

### Workflow and Output Scoring

Each prompt was inputted three times, each time by a different user (three total replicates/outputs per prompt). Two scorers independently calculated an individual score for each output to confirm consensus on all output scores; there were no discrepancies between the scorers. The final score for each prompt was calculated as an average of the three replicate scores. A schematic of the workflow can be found in Figure 1, and scoring criteria can be found in Figure 2.

**Figure 1:**
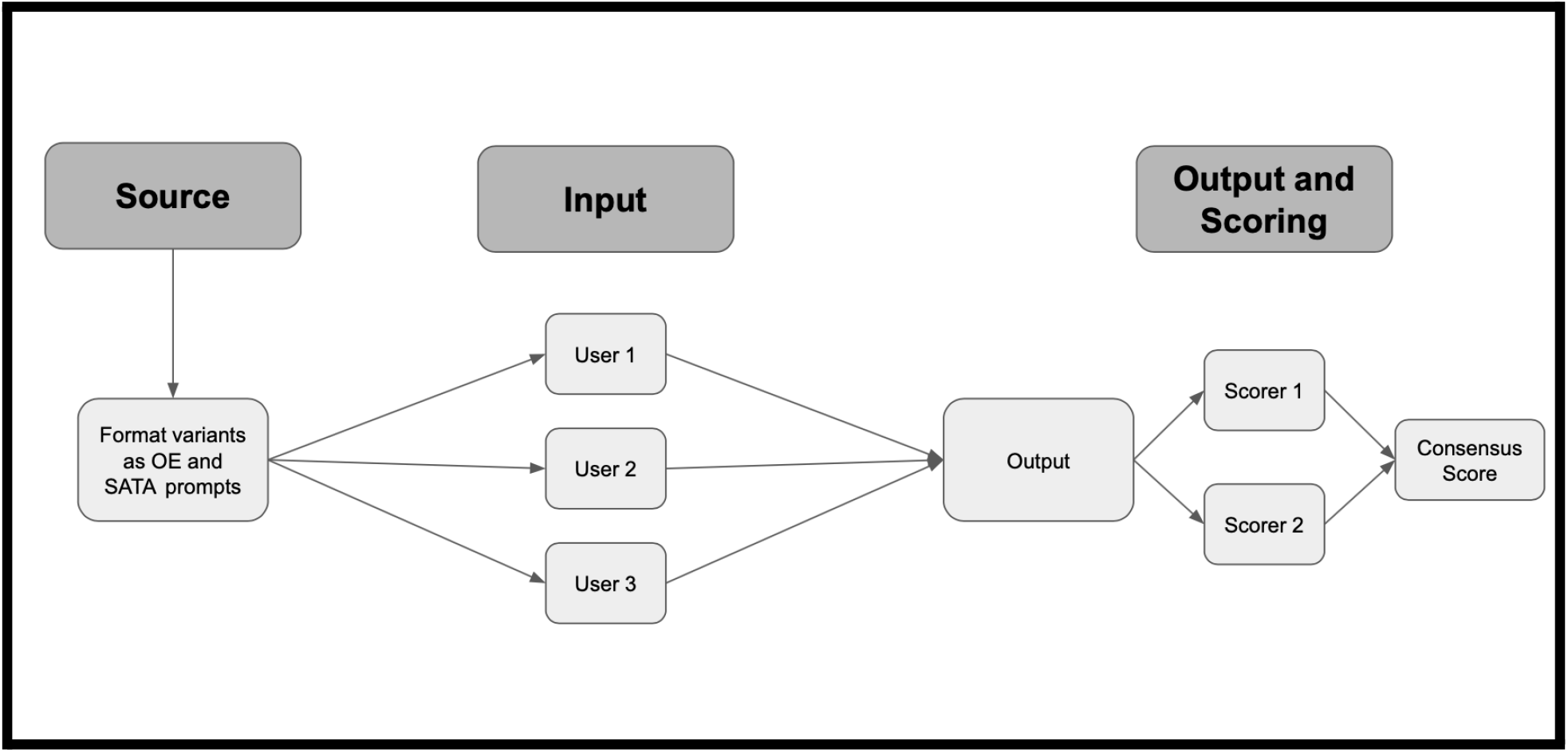
Schematic of experimental workflow. Prompts were developed from ACR variants for breast cancer screening and breast pain and converted to OE and SATA formats. Three independent users tested each prompt. Two independent scorers calculated scores for all outputs; these were compared to generate a consensus score.

**Figure 2:**
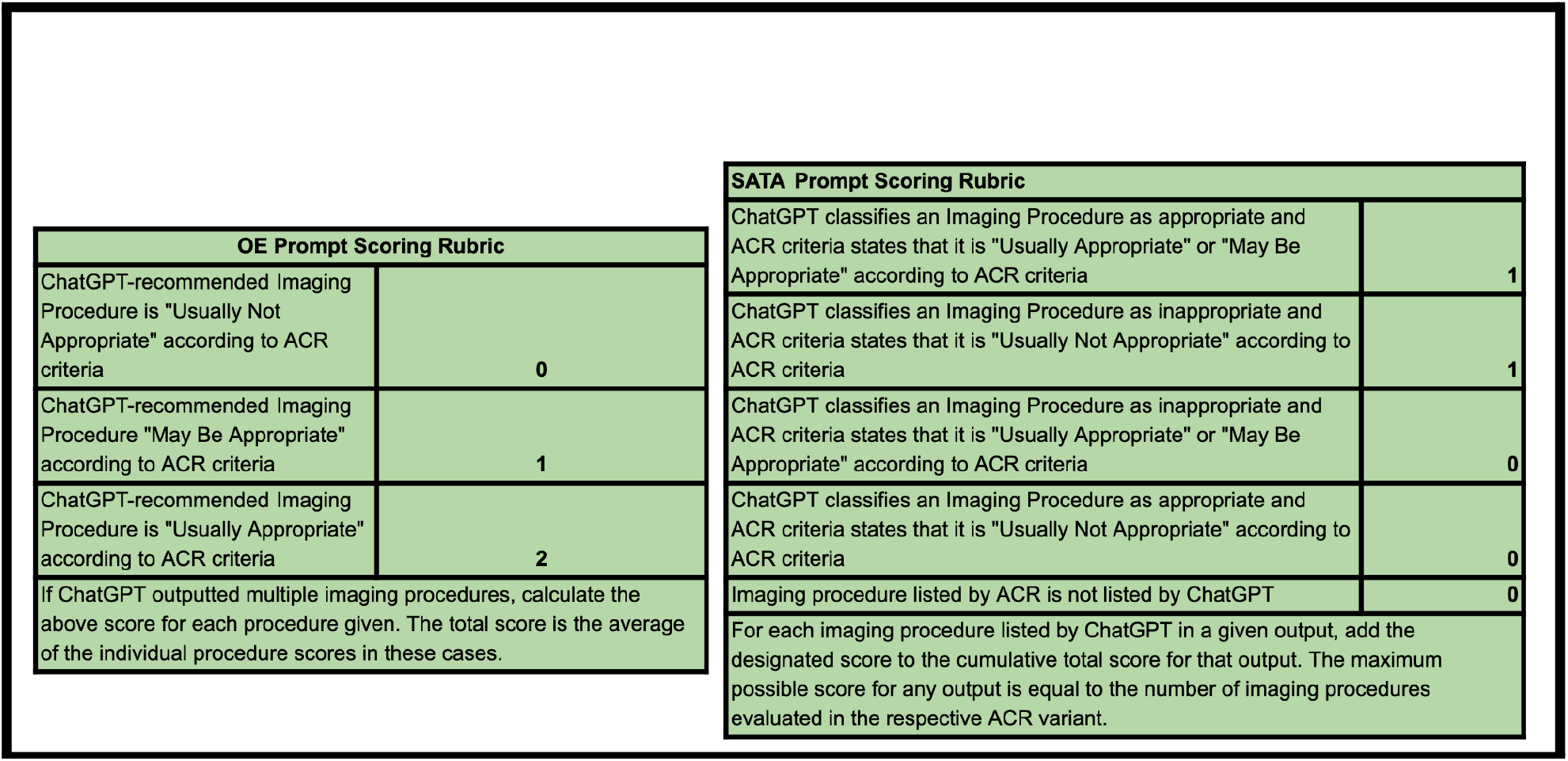
Scoring criteria for OE and SATA prompts. Answers to OE prompts were scored on a 0-2 scale, in accordance with the ACR metrics for imaging appropriateness. If multiple imaging modalities were provided for a single prompt, an individual raw score was calculated for each modality and these were averaged. Answers to SATA prompts were scored on a point/no point basis for each imaging modality provided. The maximum possible SATA score for a given variant was equal to the number of imaging procedures evaluated in the ACR criteria.

We reported the average of these raw scores for all variants. For SATA prompts, we also calculated the proportion of correct responses (the average of the raw scores divided by the maximum possible score for that variant). Since OE prompts should not yield answers which include the full spectrum of imaging options referenced by the ACR, the proportion of correct responses is not a valid metric for these outputs, thus we performed analysis on raw scores. All raw scores and additional statistical analysis can be found in the Supplementary Data.

## RESULTS

### ChatGPT achieves moderate accuracy in radiologic decision-making overall

Figures 3 and 4 show ChatGPT’s performance on all variants tested. ChatGPT shows moderate accuracy on the whole --it achieves an average OE score of 1.83 (out of 2) and a SATA average percentage correct of 88.9% for breast cancer screening prompts, and an average OE score of 1.125 (out of 2) and a SATA average percentage correct of 58.3% for breast pain prompts (Figure 3, Figure 4, Supplementary Data).

**Figure 3:**
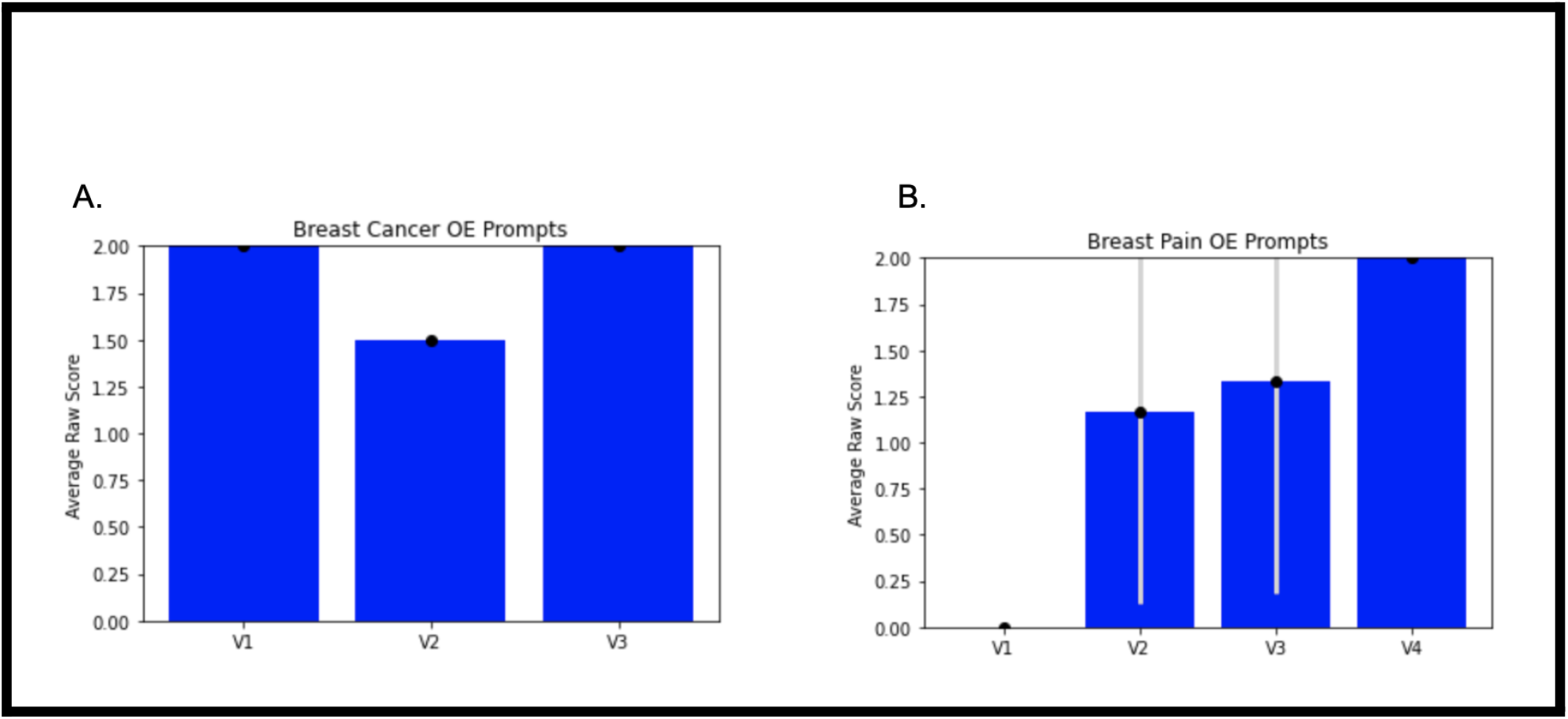
Performance of ChatGPT on OE prompts for breast cancer screening variants (Figure 3A) and breast pain variants (Figure 3B). OE performance was measured by the average raw score of the three replicate output scores for each variant (labeled according to the numbering in the ACR criteria-- “variant 1” = V1). Error bars are +/- 1 standard deviation between the three replicate output scores.

**Figure 4:**
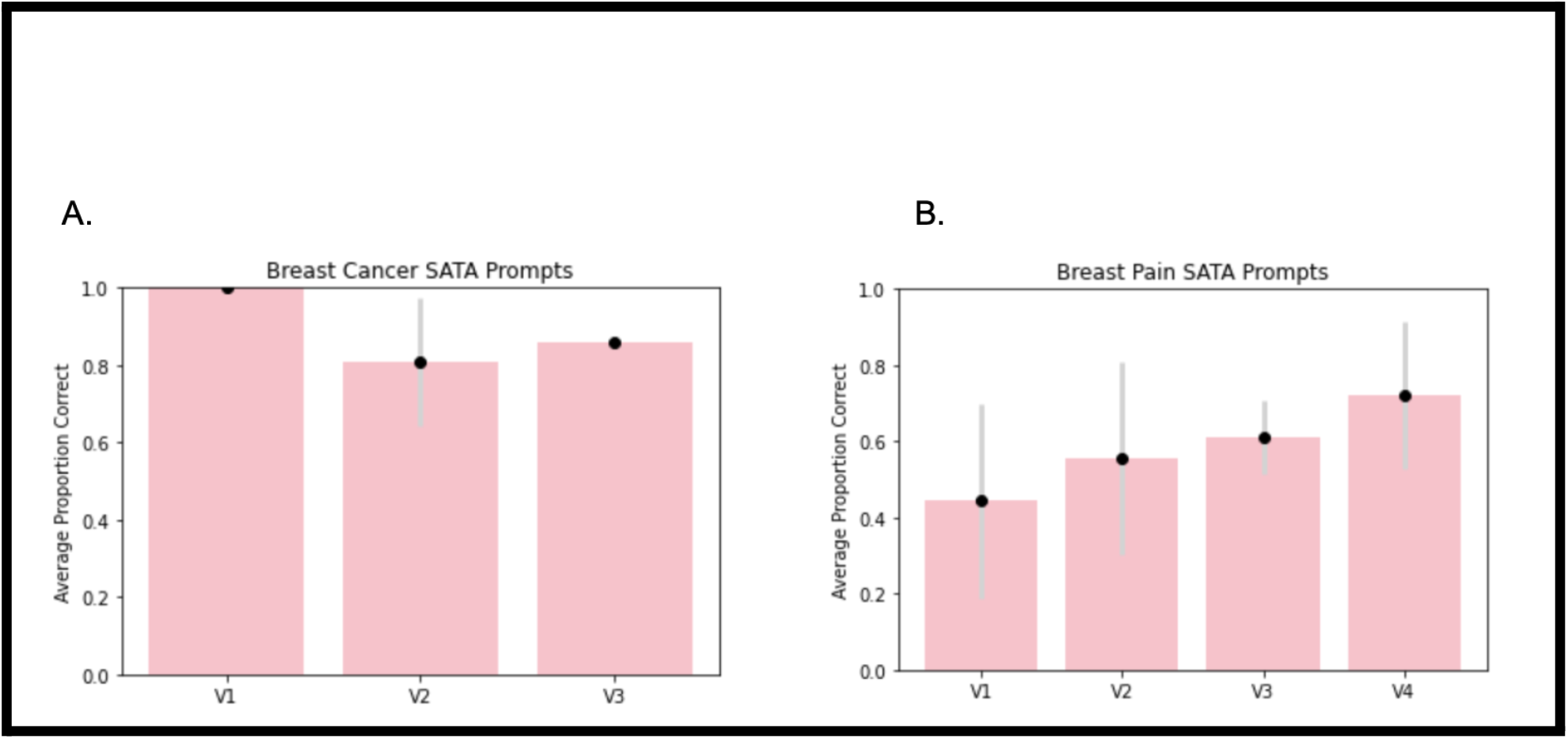
Performance of ChatGPT on SATA prompts for breast cancer screening variants (Figure 3A) and breast pain variants (Figure 3B). SATA performance was measured by the average proportion of correct answer selections for each variant from the three replicate output scores. Error bars for both prompt types are +/- 1 standard deviation between the three replicate output scores.

### ChatGPT displays more reasoning for OE prompts, but more accuracy for SATA prompts

Qualitative analysis of the raw ChatGPT output shows that OE prompts tended to yield paragraph-form answers with rationale for the choice of imaging modality. Often, such rationale would reference key points made in the ACR guidelines ^20^, such as the degree of radiation exposure. Yet, SATA responses yielded a more complete picture of the imaging considerations-- in the majority of cases, ChatGPT was not only able to identify appropriate imaging modalities, but also inappropriate imaging modalities (Figure 3, Figure 4, Supplementary Data).

### ChatGPT performance is better for breast cancer screening than breast pain

Accuracy and precision were greater for breast cancer screening prompts than for breast pain prompts, as evidenced by higher scores and lower standard deviations in the former category (Figure 3, Figure 4, Supplementary Data).

### ChatGPT accuracy varies with the severity of the initial presentation

For both the breast cancer screening and the breast pain variants, the variant number increases with the severity of the clinical presentation. For example, Variant 1 (V1) for breast cancer screening references “average-risk women”, while Variant 3 (V3) references “high-risk women” ^20^. Therefore, we were able to examine trends in ChatGPT’s triage ability. ChatGPT’s accuracy for breast cancer screening variants increased as the severity of the clinical presentation increased, while its accuracy for breast pain variants decreased as the severity increased.

### ChatGPT fails to identify futile care

Variant 1 for breast pain was the only variant of those tested whose ACR recommendation was to decline imaging services entirely. Interestingly, for both OE and SATA prompts, ChatGPT insisted on recommending imaging. Furthermore, ChatGPT provided multiple recommendations on occasion for OE prompts, even though all OE prompts asked specifically for “the single most appropriate” recommendation. These findings suggest that ChatGPT takes a maximalist approach in clinical decision-making, and is not well-equipped to identify situations in which imaging is futile.

## DISCUSSION

In this study, we provide first-of-its-kind evidence that large language models can be used as an adjunct for radiologic decision-making at the point of care. We show that ChatGPT displays moderate accuracy in determining the appropriate imaging steps for patients in need of breast cancer screening and evaluation of breast pain. Given both the intricacy of radiologic decision-making and the need for appropriate imaging utilization based only on initial clinical presentations, we believe this to be an impressive result.

With an increasingly aging population and accessible imaging technologies, radiology imaging volumes are only expected to rise despite persisting concerns for low-value imaging ^21^. Concomitant improvements in CDS infrastructure to enhance provider accuracy of appropriate imaging orders will emerge as a major priority and the involvement of ChatGPT-like AI is already being discussed ^22^.

ChatGPT performed especially well when given a set of imaging options to evaluate (Figure 3, Figure 4, Supplementary Data), consistently achieving >50% accuracy for breast pain SATA prompts and >80% for breast cancer screening SATA prompts. This is consistent with possible use cases in the clinical setting-- an ED, for example, will have a specific set of imaging modalities at its disposal, and a clinician is responsible for evaluating which of these is appropriate. ChatGPT’s performance on OE prompts was also encouraging, since it often provided extensive rationale for its recommendations, often in accordance with ACR recommendations. A hybrid approach, incorporating both a list of options for ChatGPT to evaluate and a request for ChatGPT to rationalize its choices, may provide optimal results in the clinical setting.

Most notably, ChatGPT achieved impressive accuracy for breast cancer screening prompts (on average, 88.9% correct responses for SATA prompts). Given increased efforts to reduce overutilization of imaging services in this setting ^23,24^ and the high prevalence of breast cancer in the United States ^25^, this result is especially salient.

Some important limitations of our study involve the artificial intelligence model itself. ChatGPT is not free of the inherent limitations of language models: issues of alignment with user intent (“misalignment”), fabrication of information presented (“hallucinations’’), and perhaps most arguably importantly in its potential clinical applications, inability to attribute factual information to a source. These limitations are reflected in ChatGPT’s predilection towards providing more information than requested (multiple imaging modalities when just one was requested), recommending imaging in futile situations, and providing rationale for incorrect imaging decisions. Such limitations must be considered when designing clinically-oriented prompts for use with large language models such as ChatGPT.

Because ChatGPT’s training data is not public, it is not clear whether ChatGPT was trained on the ACR criteria prior to testing. However, given that this study is only concerned with the application of existing AI tools in radiologic decision making, it is inconsequential whether or not ChatGPT was trained on ACR guidelines-- since ACR guidelines inform the standard of care, it is desirable that ChatGPT answers mirror existing guidelines, and surprising that there is not complete concordance.

As artificial intelligence-based tools and large language models specifically become more integrated with everyday use cases, we predict that specialized AI-based clinical decision-making tools will emerge. We believe that our study provides a critical data point in these endeavors, identifying the surprising strengths of AI in determining appropriate diagnostic steps and highlighting weaknesses that need to be addressed in future iterations.

## Data Availability

All data produced in the present work are contained in the manuscript

## REFERENCES

1. Bizzo, B. C., Almeida, R. R., Michalski, M. H. & Alkasab, T. K. Artificial Intelligence and Clinical Decision Support for Radiologists and Referring Providers. J. Am. Coll. Radiol. 16, 1351–1356 (2019).

2. Shah, C., Davtyan, K., Nasrallah, I., Bryan, R. N. & Mohan, S. Artificial Intelligence-Powered Clinical Decision Support and Simulation Platform for Radiology Trainee Education. J. Digit. Imaging (2022) doi:10.1007/s10278-022-00713-9.

3. Ouyang, L. et al. Training language models to follow instructions with human feedback. (2022) doi:10.48550/arXiv.2203.02155.

4. Brown, T. B. et al. Language Models are Few-Shot Learners. Preprint at https://doi.org/10.48550/arXiv.2005.14165 (2020).

5. Terwiesch, C. Would Chat GPT3 Get a Wharton MBA?

6. Bommarito, J., Bommarito, M., Katz, D. M. & Katz, J. GPT as Knowledge Worker: A Zero-Shot Evaluation of (AI)CPA Capabilities. Preprint at https://doi.org/10.48550/arXiv.2301.04408 (2023).

7. Bommarito II, M. & Katz, D. M. GPT Takes the Bar Exam. Preprint at https://doi.org/10.48550/arXiv.2212.14402 (2022).

8. Kung, T. H. et al. Performance of ChatGPT on USMLE: Potential for AI-Assisted Medical Education Using Large Language Models. 2022.12.19.22283643 Preprint at https://doi.org/10.1101/2022.12.19.22283643 (2022).

9. Stokel-Walker, C. ChatGPT listed as author on research papers: many scientists disapprove. Nature 613, 620–621 (2023).

10. Biswas, S. ChatGPT and the Future of Medical Writing. Radiology 223312 (2023) doi:10.1148/radiol.223312.

11. Flanagin, A., Bibbins-Domingo, K., Berkwits, M. & Christiansen, S. L. Nonhuman “Authors” and Implications for the Integrity of Scientific Publication and Medical Knowledge. JAMA (2023) doi:10.1001/jama.2023.1344.

12. Thorp, H. H. ChatGPT is fun, but not an author. Science 379, 313–313 (2023).

13. Tools such as ChatGPT threaten transparent science; here are our ground rules for their use. Nature 613, 612–612 (2023).

14. Sadigh, G. et al. Downstream Breast Imaging Following Screening Mammography in Medicare Patients with Advanced Cancer: A Population-Based Study. J. Gen. Intern. Med. 33, 284–290 (2018).

15. Schonberg, M. A. Overutilization of Breast Cancer Screening in the US: Awareness of a Growing Problem. J. Gen. Intern. Med. 33, 238–240 (2018).

16. Habib, A. R., Grady, D. & Redberg, R. F. Recommendations From Breast Cancer Centers for Frequent Screening Mammography in Younger Women May Do More Harm Than Good. JAMA Intern. Med. 181, 588–589 (2021).

17. Goyal, A. Breast pain. BMJ Clin. Evid. 2014, 0812 (2014).

18. Kushwaha, A. C. et al. Overutilization of Health Care Resources for Breast Pain. Am. J. Roentgenol. 211, 217–223 (2018).

19. ChatGPT General FAQ. https://help.openai.com/en/articles/6783457-chatgpt-general-faq.

20. ACR Appropriateness Criteria®. https://www.acr.org/Clinical-Resources/ACR-Appropriateness-Criteria.

21. Kjelle, E. et al. Characterizing and quantifying low-value diagnostic imaging internationally: a scoping review. BMC Med. Imaging 22, 73 (2022).

22. Shen, Y. et al. ChatGPT and Other Large Language Models Are Double-edged Swords. Radiology 230163 (2023) doi:10.1148/radiol.230163.

23. Sharma, R. et al. Factors Influencing Overuse of Breast Cancer Screening: A Systematic Review. J. Womens Health 27, 1142–1151 (2018).

24. Austin, J. D. et al. A mixed-methods study of multi-level factors influencing mammography overuse among an older ethnically diverse screening population: implications for de-implementation. Implement. Sci. Commun. 2, 110 (2021).

25. Giaquinto, A. N. et al. Breast Cancer Statistics, 2022. CA. Cancer J. Clin. 72, 524–541 (2022).

